# The timing of surgical interventions following the implantation of coronary drug-eluting stents in patients undergoing gastrointestinal cancer surgery: A multicenter retrospective cohort study

**DOI:** 10.1101/2024.02.05.24302371

**Authors:** Ziyao Xu, Xinyu Hao, Jingyang Tian, Qiying Song, Tian Li, Lei Gao, Xinxin Wang

## Abstract

**Background:** The guidelines recommending a minimum deferral of six months for non-cardiac surgeries following drug-eluting stent percutaneous coronary intervention (DES-PCI) do not adequately address the requirements for individuals undergoing gastrointestinal cancer surgery (GCS). We aim to investigate the optimal timing for surgical interventions to maximize patient benefit.

**Methods:** The study encompassed 2,501 patients treated from January 2017 to December 2021, all of whom underwent GCS within one year after DES-PCI. We conducted an analysis by comparing the occurrence of Major Adverse Cardiovascular Events (MACEs) within 30 days post-surgery at different time points.

**Results:** This study enrolled a total of 2501 participants with meticulously recorded data who underwent DES-PCI and subsequently underwent GCS within one year post-implantation. The incidence rate of MACEs is 14.2%, including MI(5.1%), HF(5.8%), IS(3.2%), Cardiac death(0.2%) across all patients in this study. The threshold probability was determined using the Youden Index, resulting in a value of 0.320, corresponding to a “Time of surgery value” of 87. Significant statistical differences were observed in the occurrence rates of MACEs for adjacent time intervals at 30 days (p < 0.001), 90 days (p < 0.009), and 180 days (p < 0.001).

**Conclusions:** The timing of surgical intervention following DES-PCI significantly influences the occurrence of MACEs at 1 month, 3 months, and 6 months. GCS may be appropriately advanced within the 6-month timeframe, but with the exception of emergency, efforts should be made to defer them beyond the initial month.

## 1. Introduction

In patients with coronary stents, the probability of undergoing various surgeries within one year is approximately 4% to 20%, with around 86% involving non-cardiac surgeries (NCS)[1]. Guidelines recommend that all patients should undergo elective NCS at least 6 months after drug-eluting stent percutaneous coronary intervention (DES-PCI). However, if the risk of delaying surgery is higher, consideration may be given to performing the surgery 3 to 6 months after DES implantation, with uninterrupted dual antiplatelet therapy (DAPT) during the postponement period[2]. Notably, in patients with a history of stent implantation undergoing gastrointestinal cancer surgery (GCS), up to 7.5% may require surgery within 6 months after DES-PCI[3]. This may be attributed to the bleeding tendency and difficulty in eating associated with GCS, making the surgery more urgent[4].

In surgical procedures for GCS, lymph node dissection, vascular ligation, and reconstruction of the digestive tract are essential components[5]. Due to variations in tumor staging, combined organ resection may be necessary, rendering digestive tract tumor cancer surgery associated with a higher risk of bleeding compared to other NCS[6–8]. Additionally, prematurely undergoing surgery for digestive tract cancer after coronary stent placement may lead to increased occurrence of major adverse cardiovascular events (MACEs) postoperatively[9]. Therefore, the timing of digestive tract tumor surgery after coronary stent implantation is crucial, representing a delicate balance between ischemic and bleeding risks during the perioperative period[10–12]. This ultimately determines whether patients can benefit from undergoing surgical intervention earlier[13]. Currently, there exists no definitive guideline elucidating the optimal timing for NCS following DES-PCI. This study aims to explore the postoperative risk of MACEs in patients undergoing digestive tract cancer surgery at different time intervals after coronary stent implantation, with the goal of investigating the feasibility of advancing the surgical timing for this patient population.

## 2. Methods

### 2.1 Patients and design

We conducted this study using multicenter data from three independent hospitals, including the First Medical Center of the PLA General Hospital, the Sixth Medical Center of the PLA General Hospital, and the Hainan Hospital of the PLA General Hospital. The study encompassed 2,501 patients treated from January 2017 to December 2021, all of whom underwent gastrointestinal cancer surgery within one year after coronary stents implantation.

This research received approval from the Ethics Committee of the PLA General Hospital in China, with approval number S2023-630, exempting patients from informed consent.

### 2.2 Inclusion and exclusion criteria

Patients with gastrointestinal cancer post-coronary stent implantation were included in the study, classified according to the 8th edition of the American Joint Committee on Cancer (AJCC) TNM staging criteria as T1∼4N1∼3M0. Exclusion criteria were as follows: (1) Presence of tumors in other systems. (2) Concurrent presence of organic heart disease other than coronary artery disease. (3) History of previous cardiac vascular bypass grafting. (4) Receipt of neoadjuvant chemotherapy. (5) Unclear documentation of the time interval between stent implantation and gastrointestinal cancer surgery.

### 2.3 Outcomes

We enlisted attending physicians with extensive clinical experience to identify major adverse cardiovascular events (MACEs) in patients as the primary outcomes for this study. The endpoint was defined as the occurrence of myocardial infarction, ischemic stroke, heart failure, and cardiovascular death within 30 days following gastrointestinal cancer surgery[14].

We employed a recognized healthcare system capable of structured storage and extraction, retrieving relevant clinical data by searching sensitive word fields from the clinical treatment digital system’s corpus to determine clinical events. The specialized application of this system overcame difficulties in identifying different clinical events across diverse data fields. MACEs were diagnosed by retrieving patient medical records, prescription records, nursing records, laboratory test results, ECGs, and echocardiograms. Each of the three independent medical centers recruited three senior physicians for collaborative diagnosis of complications, resolving disputed matters through debate and consensus.

In this study, the diagnosis of myocardial infarction (MI) requires alignment with clinical symptoms of ischemic chest pain and concomitant changes in cardiac biomarker laboratory assays. Primarily, there should be an elevation of myocardial markers exceeding the 99th percentile of the upper reference limit with dynamic changes. Additionally, at least one of the following four criteria must be met: (a) symptoms of chest tightness or angina; (b) electrocardiogram indicating ischemic changes; (c) presence of pathological Q waves; (d) identification of coronary artery thrombus during percutaneous coronary intervention (PCI) or post-mortem autopsy[15]. IS is a disease associated with vascular obstruction, characterized by sudden onset symptoms such as facial paralysis, speech impediments, and limb weakness[16]. Confirmation of ischemic damage is typically achieved through neuroimaging techniques such as Magnetic Resonance Imaging (MRI) or Computed Tomography (CT) scans[17]**^Error!^ ^Reference^ ^source^ ^not^ ^found.^**. HF is characterized by the inability to effectively pump blood, leading to impaired blood circulation. Typical symptoms include shortness of breath, fatigue, edema (especially in the lower extremities), and palpitations. Diagnosis often requires cardiac imaging and laboratory tests to assess cardiac structure and function[18]. Cardiac death refers to mortality occurring in the natural course of cardiac pathology, rather than being attributed to external factors. It is associated with severe cardiac conditions or significant abnormalities in the cardiovascular system, often accompanied by symptoms such as severe heart failure, arrhythmias, myocardial infarction[19].

### 2.5 Statistical analysis

In the analysis, categorical variables were expressed as frequencies and percentages. For continuous variables following a normal distribution, descriptive statistics included the mean and standard deviation, while those deviating from normality were characterized by the median and interquartile range (IQR). To assess the normality of the data, the Kolmogorov-Smirnov test was employed. Subsequently, for normally distributed continuous variables, either a t-test or ANOVA was applied, depending on the nature of the comparison. In cases where normal distribution assumptions were violated, the analysis utilized the Whitney U test or Kruskal-Wallis test. Similarly, the examination of categorical variables involved the use of either the chi-square test or Fisher’s exact test to assess the correlation and significance among different groups.

Building upon the aforementioned statistical methods for inter-group data analysis, we employed a multifactorial logistic regression analysis to elucidate the impact of different variables on the outcome, concurrently ranking the importance of these variables. Based on this foundation, we established a logistic regression equation for the variable "Time of surgery" and its association with the outcome of Major Adverse Cardiovascular Events (MACEs). The logistic regression equation was constructed using the Jordan index to compute the threshold probability, ultimately yielding specific numerical values corresponding to the surgical timing associated with this threshold probability.

We utilized the ggplot2 package in Rstudio to create a bar chart illustrating the relationship between surgical timing and MACEs. Statistical significance between different bars was assessed using chi-square tests and error bars. In accordance with the ranking of variable importance, a descriptive analysis of the relationship between time and emergency with MACEs was conducted. Additionally, a line density plot, considering emergency as a variable, was generated to reflect the number of emergency surgeries at different surgical timings. Finally, Kaplan-Meier curves were plotted to depict the trend variations in the occurrence of MACEs for different surgical timings, with statistical significance between surgical timing intervals compared through Log-rank tests. The aforementioned visualization procedures were consistently executed using the ggplot2 package.

Data were statistically analyzed using R statistical software (R version 4.2.3, R Foundation for Statistical Computing). Statistical significance was accepted at the 0.05 level, and all tests were two-tailed.

## 3. Results

### 3.1 Baseline of clinical data

This study enrolled a total of 2501 participants with meticulously recorded data who underwent DES-PCI and subsequently underwent GCS within one year post-implantation. The multicenter data collection was conducted at the First Medical Center of the Chinese People’s Liberation Army General Hospital, the Sixth Medical Center of the Chinese People’s Liberation Army General Hospital, and the Hainan Hospital of the Chinese People’s Liberation Army General Hospital, adhering to the aforementioned inclusion criteria. Among these participants, 354 cases experienced MACEs. The detailed procedures for patient screening and recruitment are illustrated in Figure 1.

**Figure 1.**
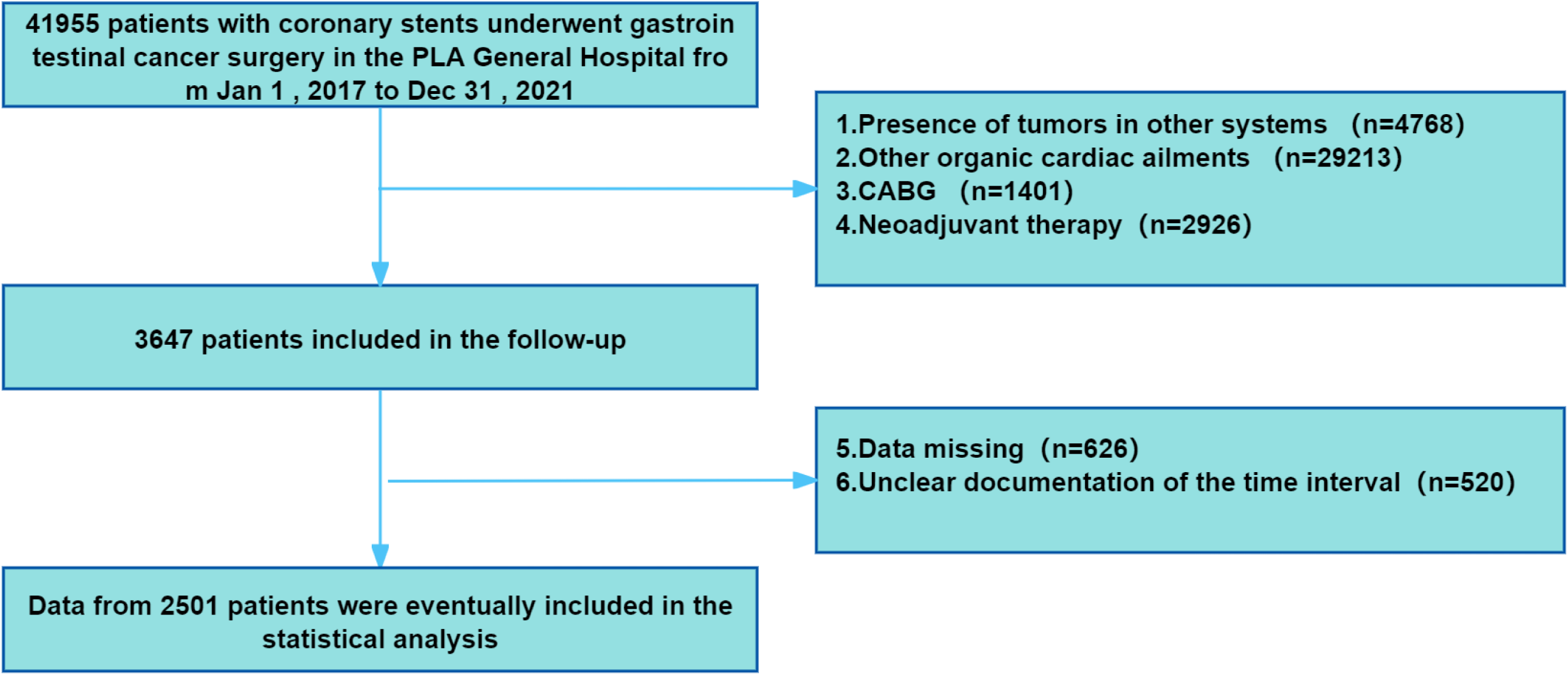
Flowchart of the study. CABG, coronary artery bypass grafting.

Table 1 encapsulates baseline data, encompassing demographic parameters, medical history, medication records, clinical-pathological staging of tumors, cardiac-related variables, preoperative laboratory indicators, and surgical variables. The data were stratified based on the presence or absence of MACEs, and inter-group disparities were scrutinized.

**Table 1.**
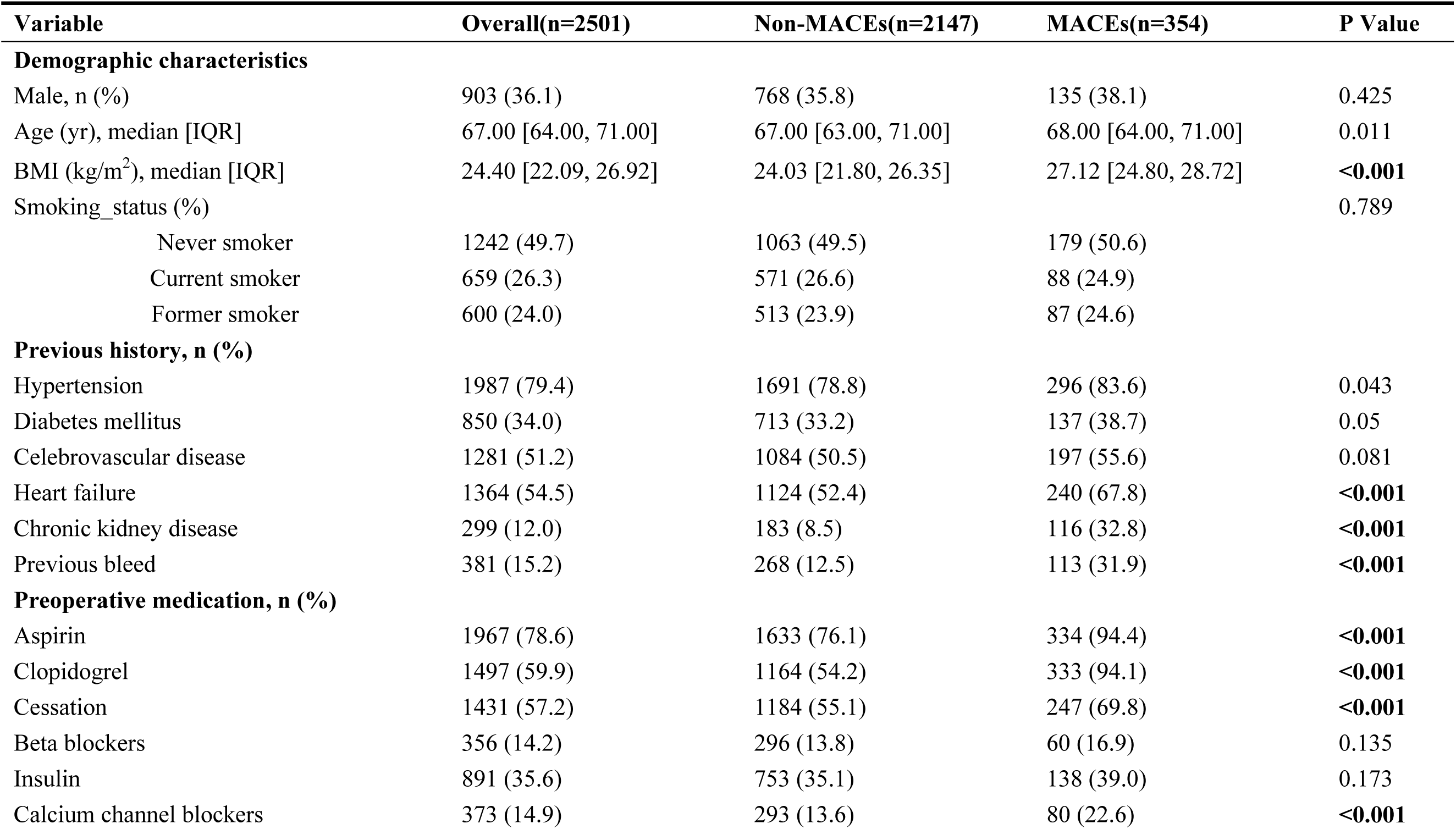
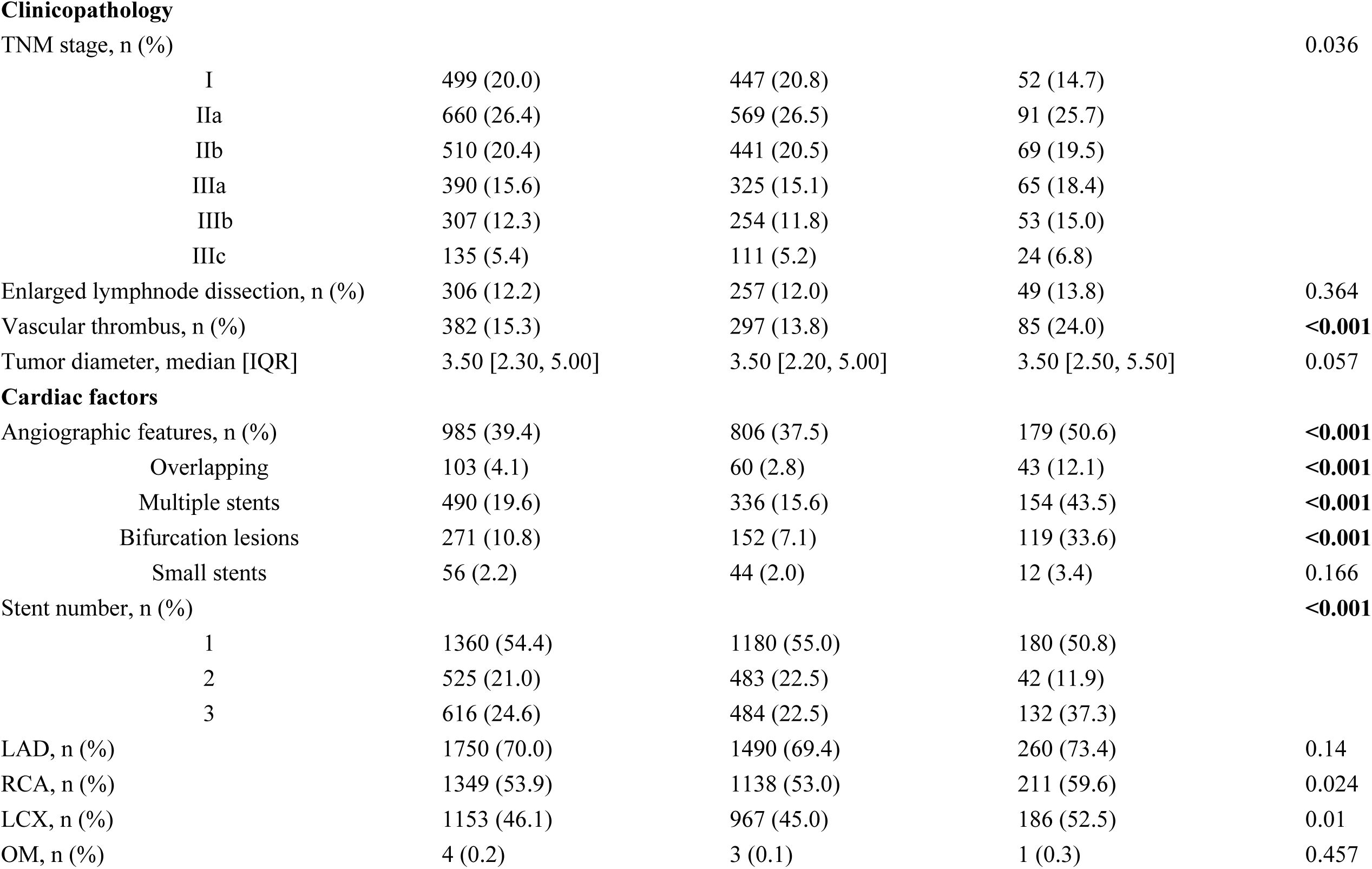
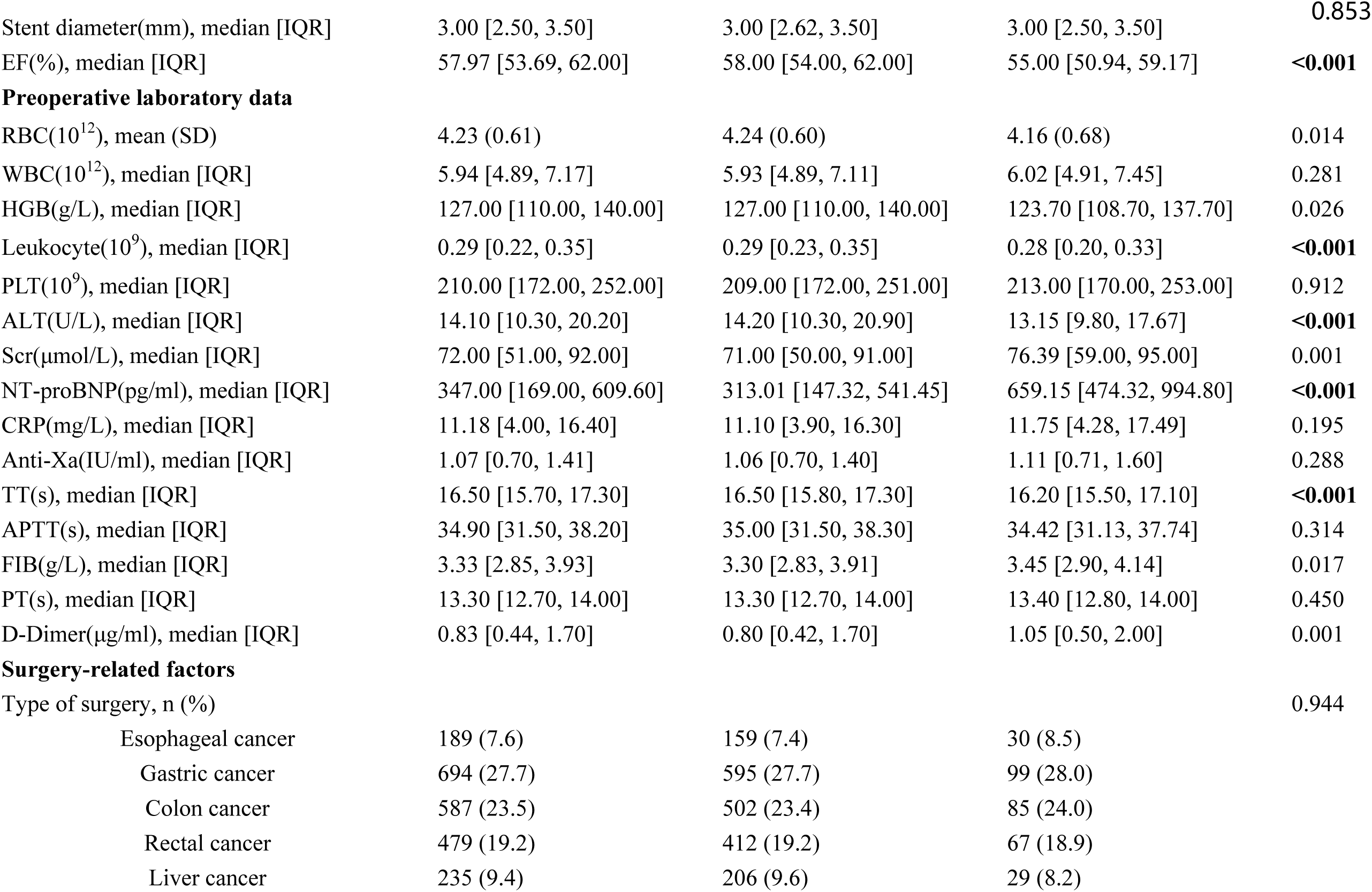
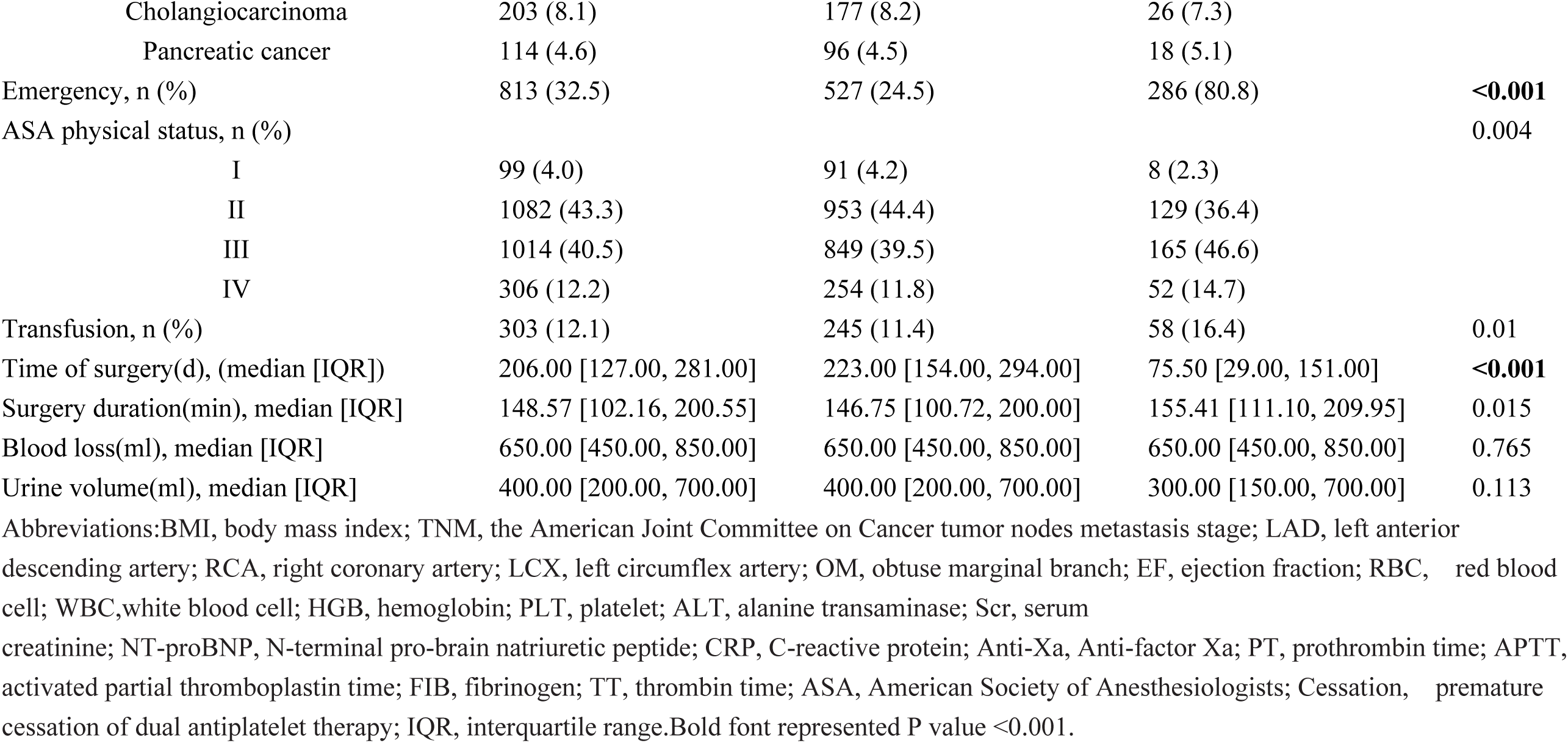
Baseline characteristics of the patients with and without MACEs.

In the realm of demographic characteristics, only the BMI of the MACEs group exhibited a significant elevation compared to the non-MACEs group. Within medical history, the MACEs group displayed a substantial increase in the prevalence of heart failure (67.8%), chronic kidney disease (32.8%), and prior bleeding episodes (31.9%) relative to the non-MACEs group. In the context of medication history, a noteworthy surge was observed in the MACEs group concerning aspirin (94.4%), clopidogrel (94.1%), calcium channel blockers (22.6%), and premature termination of antiplatelet medications (69.8%) compared to the non-MACEs group.

Analysis of the pathological staging of gastrointestinal tumors revealed no significant distinctions between the MACEs and non-MACEs groups. Regarding cardiac-related variables, patients in the MACEs group demonstrated a markedly higher incidence of overlapping stents (12.1%), multiple stents (43.5%), and multiple lesions (33.6%) than those in the non-MACEs group. Additionally, the ejection fraction (EF) was notably lower in the MACEs group (55.00% [50.94, 59.17]) in contrast to the non-MACEs group (58.00% [54.00, 62.00]).

Within the realm of surgical variables, the frequency of emergency surgeries was significantly higher in the MACEs group (80.8%), while the interval between surgeries was significantly shorter (75.50 days [29.00, 151.00]) compared to the non-MACEs group. Figure 2 depicts the occurrence rates of MACEs(14.2%), including MI(5.1%), HF(5.8%), IS(3.2%), Cardiac death(0.2%) across all patients in this study.

**Figure 2.**
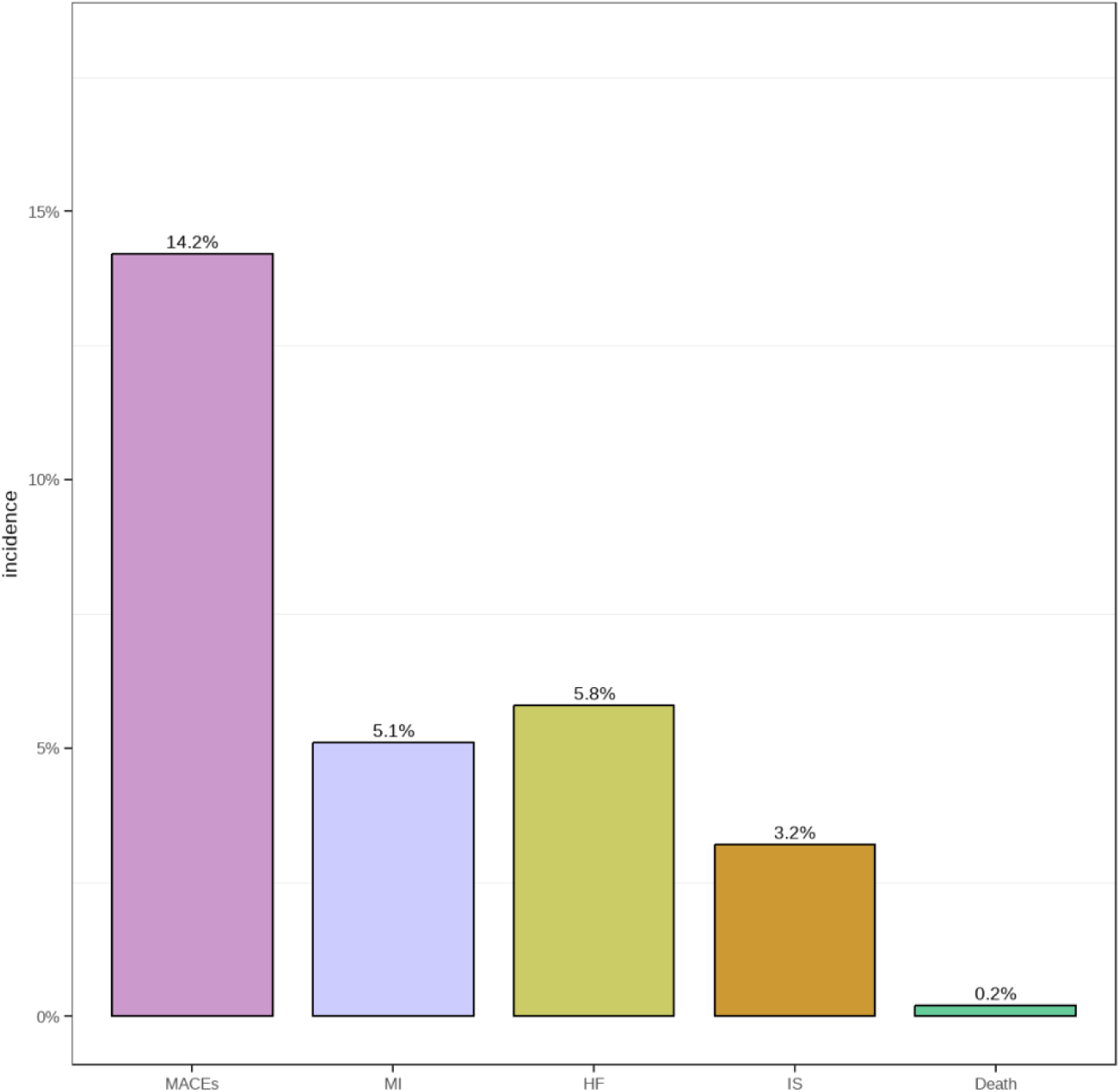
Comparison of the proportion of patients with MACEs. MACEs, major adverse cardiovascular events; MI, non-fatal myocardial infarction, IS, ischemic stroke, HF,heart failure

### 3.2 The logistic regression analysis on MACEs

Table 2 presents the results of a multivariable logistic regression analysis on the outcome of MACEs, following single-factor analysis based on Table1. The analysis includes the incorporation of eleven variables into the model: Time of surgery, emergency, Heart failure, Hypertension, Chronic kidney disease, Body mass index (BMI), Clopidogrel, Vascular thrombus, Overlapping, Ejection fraction (EF), and N-terminal pro-brain natriuretic peptide (NT-proBNP), highlighting them as independent risk factors for MACEs. Additionally, insights into the importance ranking of variables influencing MACEs from the multivariable logistic regression can be derived from Figure 3. Notably, Time of surgery and emergency exhibit a pronounced impact on MACEs.

**Figure 3.**
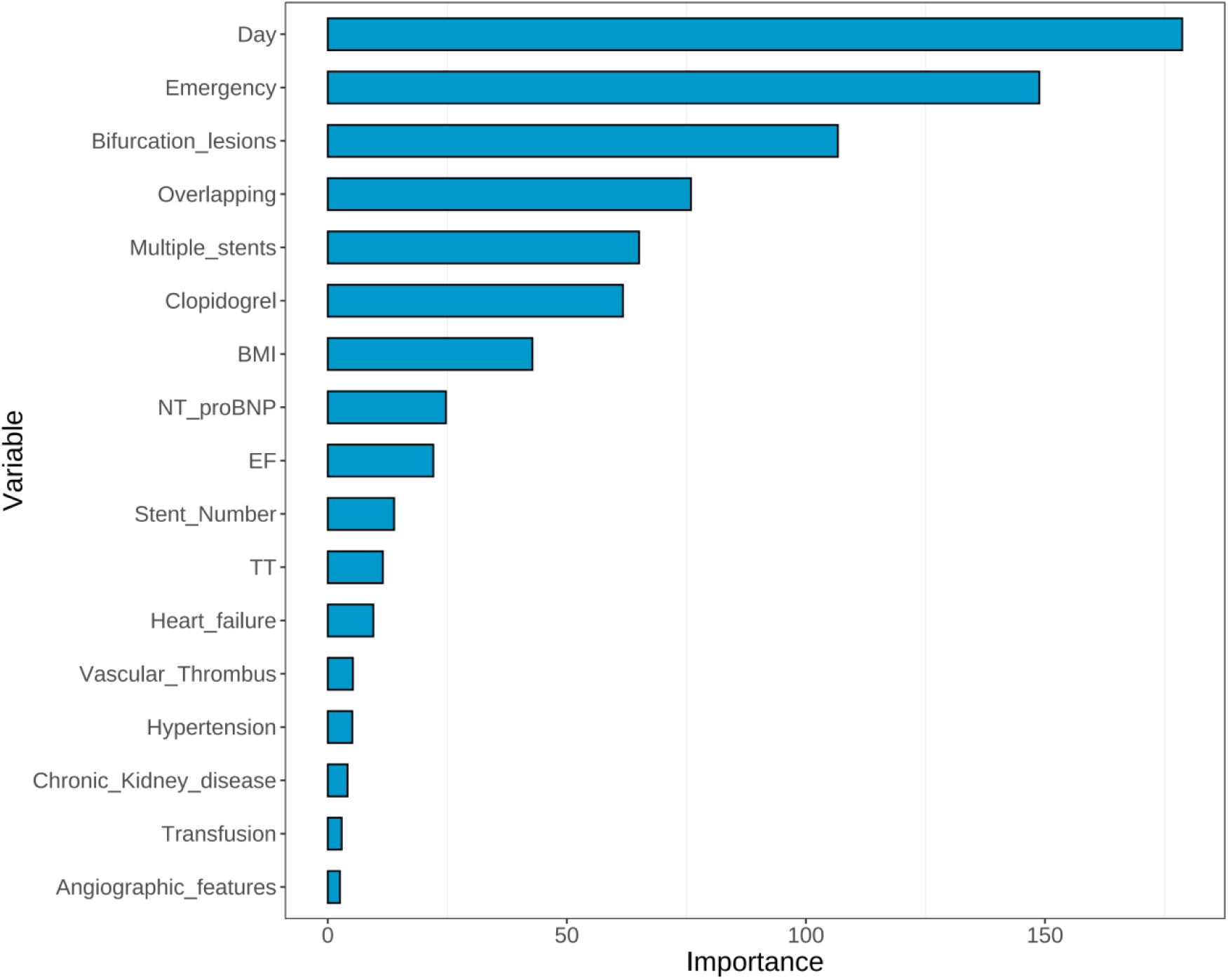
Importance ranking of variables affecting MACEs. BMI, body mass index; EF, ejection fraction; TT, thrombin time.

**Table 2.** Multivariate logistic regression analysis.

| Variable | Multivariate analysis |  |  |
| --- | --- | --- | --- |
|  | B | OR (95% CI) | P value |
| Heart failure | 0.670 | 1.954(1.270–3.007) | **<0.001** |
| Hypertension | 0.645 | 1.906(1.076-3.374) | **<0.001** |
| Chronic kidney disease | 0.550 | 1.734(1.021-2.946) | **<0.001** |
| BMI | 0.223 | 1.249(1.165-1.340) | **<0.001** |
| Clopidogrel | 2.118 | 8.314(4.574–15.112) | **<0.001** |
| Vascular thrombus | 0.628 | 1.873(1.100-3.192) | **<0.001** |
| Overlapping | 0.339 | 8.083(5.827–13.634) | **<0.001** |
| EF | -0.077 | 0.926(0.896–0.957) | **<0.001** |
| NT-proBNP | 0.012 | 1.004(1.001-1.010) | **<0.001** |
| Emergency | 2.539 | 12.673(8.051-19.947) | **<0.001** |
| Time of surgery | -0.518 | 0.986(0.983-0.988) | **<0.001** |

Abbreviations: BMI, body mass index; EF, ejection fraction; NT-proBNP, N-terminal pro-brain natriuretic peptide. Bold font represented P value <0.05.

### 3.3 The Association between the pivotal variables “Time of surgery and Emergency” and MACEs

From Figure 4A, it is evident that the incidence rate of MACEs in emergency surgeries is significantly higher than that in elective surgeries. Moreover, with an increasing time interval between DES-PCI and GCS, the MACEs incidence rates for both exhibit a declining trend. Additionally, considering the surgical volume, the daily emergency surgeries for gastrointestinal cancer are higher than elective surgeries when the time interval between coronary stent placement and gastrointestinal cancer surgery ranges from 0 to 180 days. Furthermore, within the time interval of 0-30 days, the surgical volume for emergency gastrointestinal surgeries rapidly increases, reaching its peak at the thirtieth day (Figure 4B).

**Figure 4.**
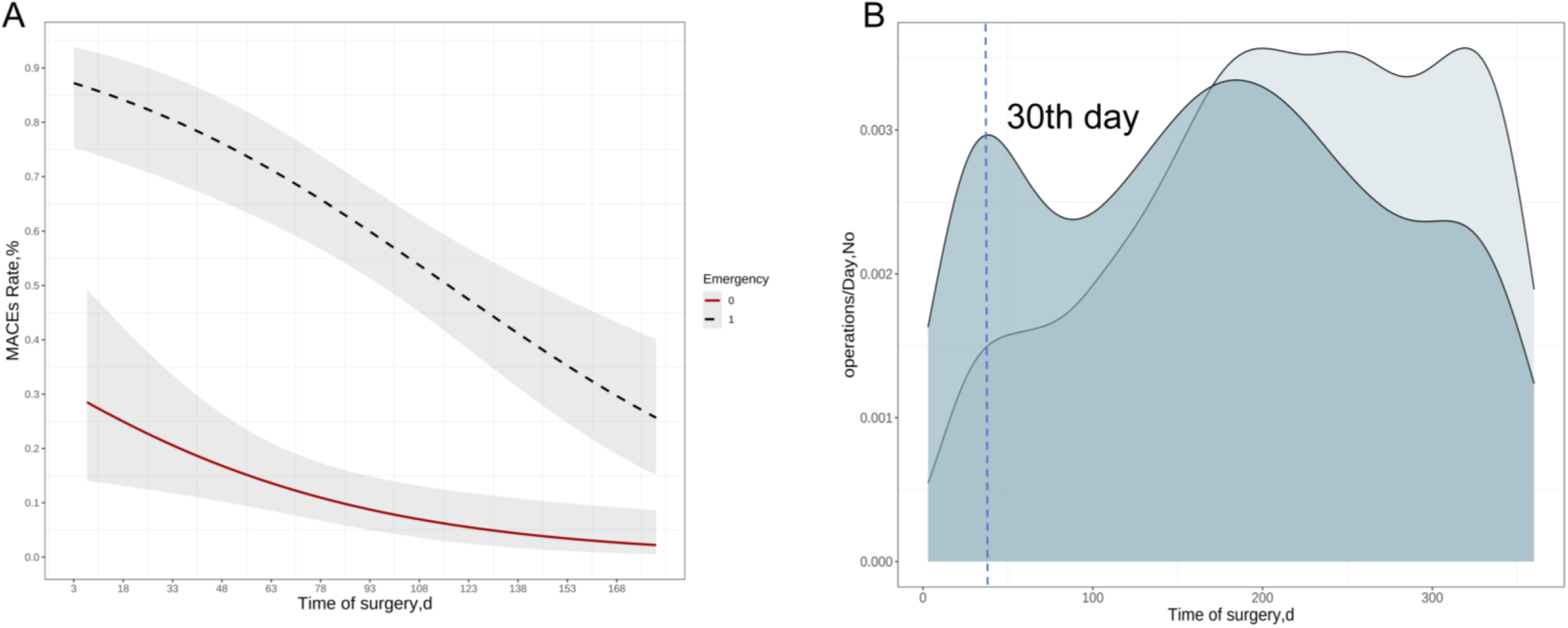
The trend of emergency and elective surgery with changes in surgical timing. (A) Line chart reflecting the trend of changes in MACEs after emergency and elective surgery. (B) Surgical volume density map of emergency and elective surgery at different time.

### 3.4 Comparison of the incidence rates of MACEs at different surgical timings

Basing our analysis on the logistic regression equation of “Time of surgery and MACEs”, the threshold probability was determined using the Youden Index, resulting in a value of 0.320, corresponding to a “Time of surgery value” of 87 (refer to Supplementary Material 1 for detailed calculations). Additionally, dividing the surgical time interval into consecutive 30-day intervals, significant statistical differences were observed in the occurrence rates of MACEs for adjacent time intervals at 30 days (p < 0.001), 90 days (p < 0.009), and 180 days (p < 0.001, Figure 5). Furthermore, the incidence rates of MACEs for all patients decrease with an extended surgical interval. Moreover, within the 0-30 days interval, 30-90 days interval, and 90-360 days interval, the survival rates of MACEs exhibit a stepwise increase. Significantly, through Log-Rank tests, statistically significant differences were observed between any two groups (Figure 6).

**Figure 5.**
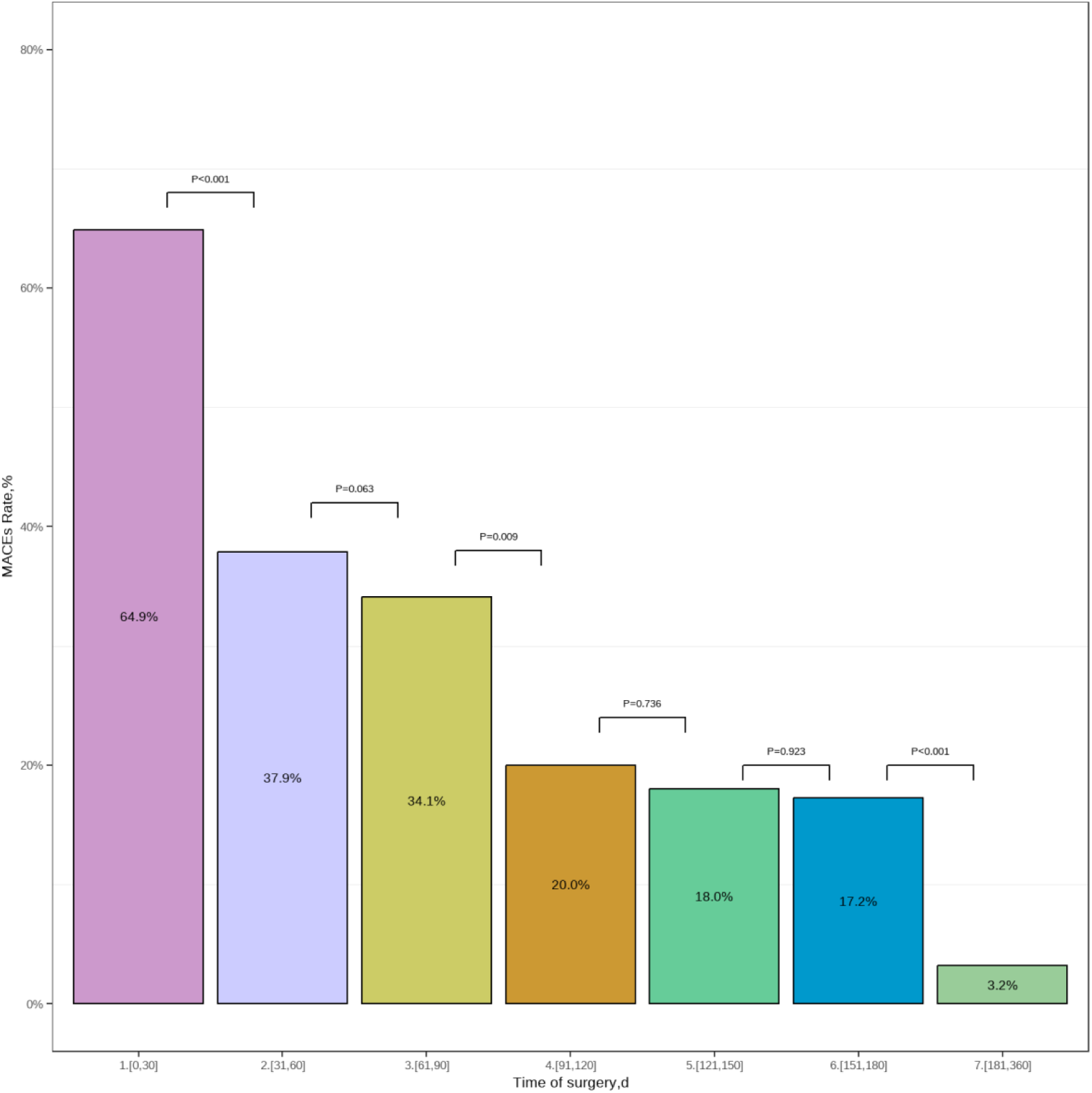
A bar chart with error bars comparing the occurrence rates of MACEs across different time intervals.

**Figure 6.**
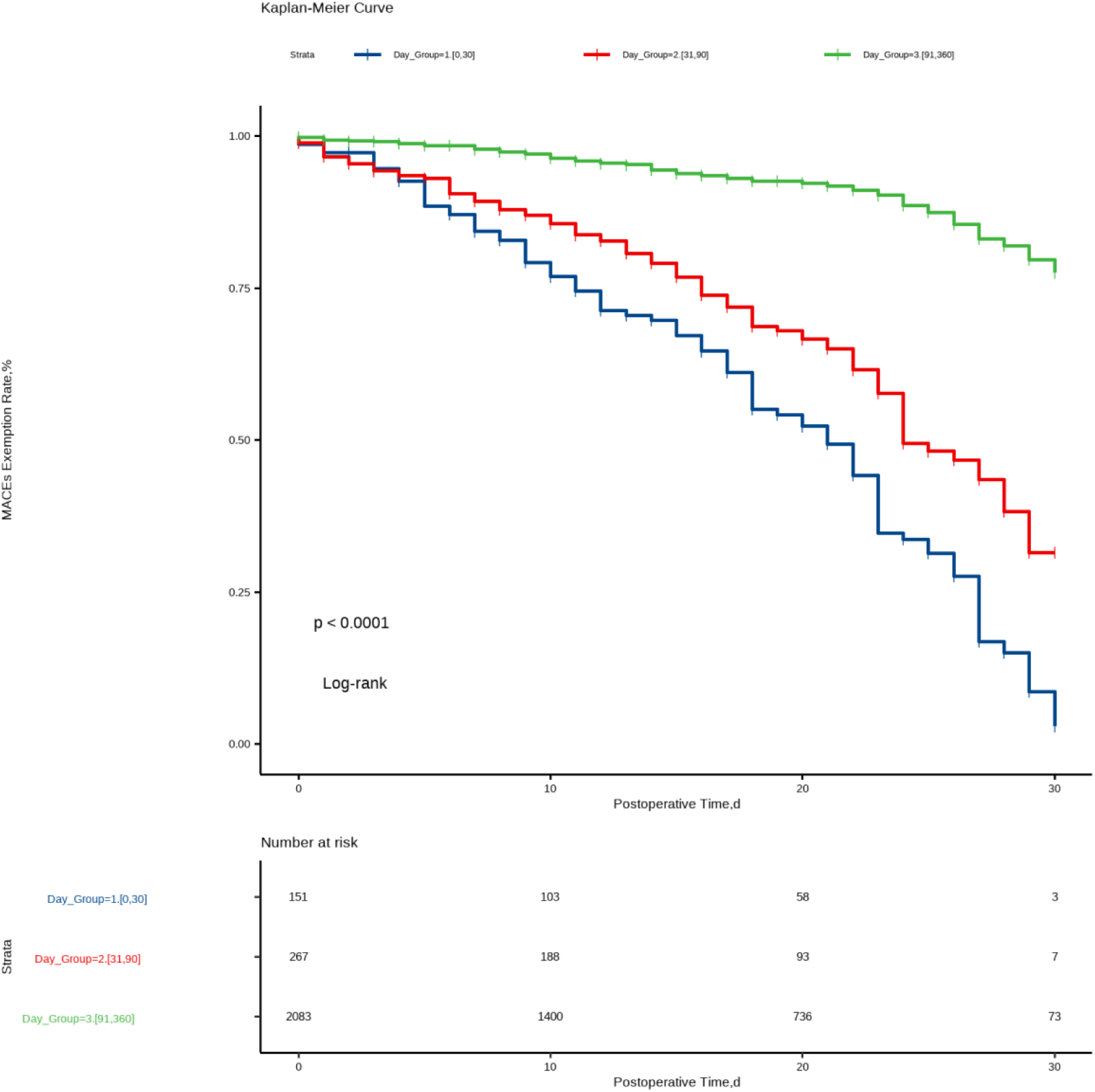
K-M curve of 30-day postoperative MACE exemption rate for patients grouped at different surgical timing.

**Figure 7.**
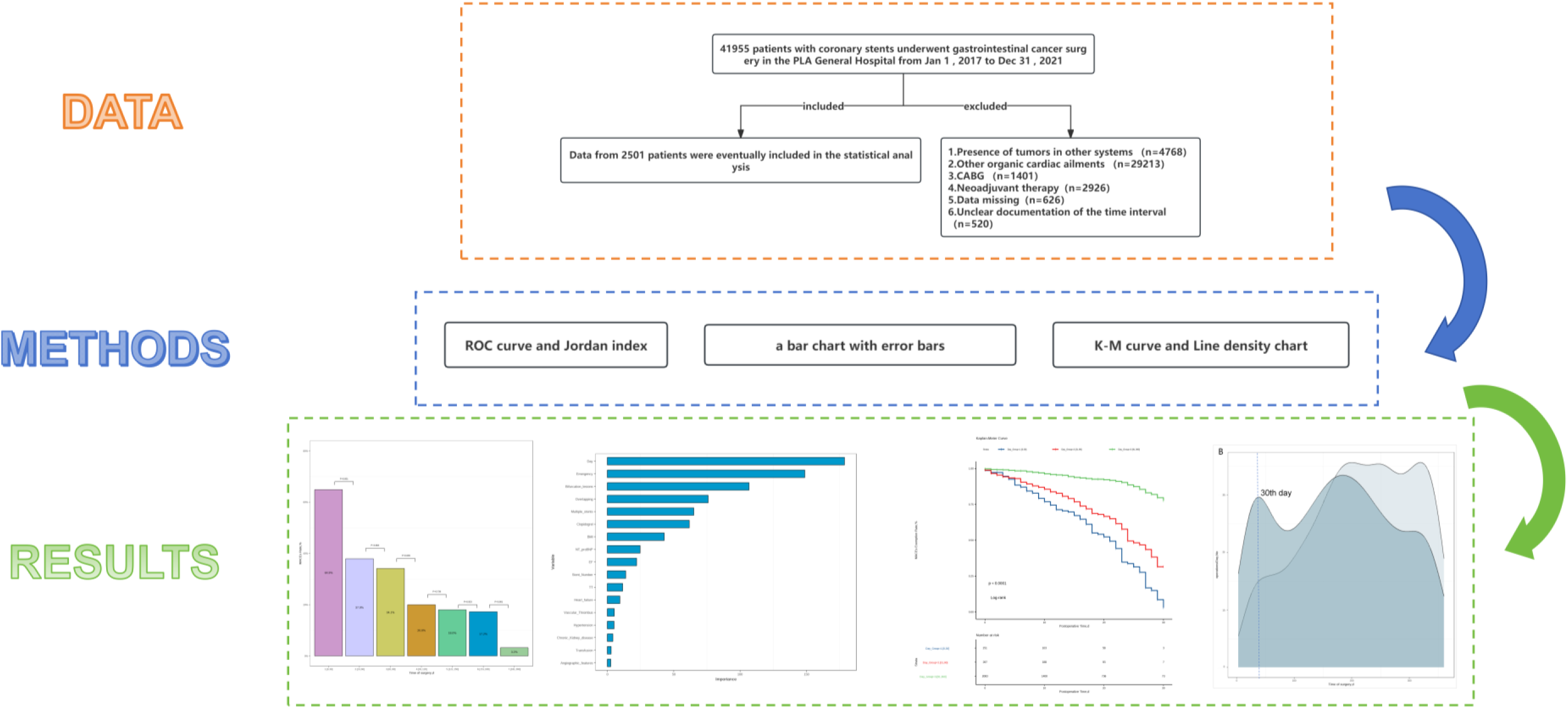
Exploring the optimal surgical timing for patients undergoing DES-PCI.

## 4. Discussion

### 4.1 Main interpretation

In this study, we utilized data from multicenter medical records to investigate the occurrence rates of MACEs within 30 days after GCS in patients who underwent coronary stent implantation. Through comparative analyses across different time intervals, the following key findings emerged after employing various statistical approaches:(1) Significant statistical differences in MACEs occurrence rates were observed between adjacent time intervals, namely 0-30 days (63%), 30-60 days (38%), 60-90 days (35%), 90-120 days (20%), 150-180 days (17%), and 180-360 days (2%). (2) The MACEs exemption rates within the consecutive time intervals of 0-30 days, 30-90 days, and 90-360 days exhibit a progressive decline over time after surgery. Importantly, the statistical disparities between adjacent intervals are deemed significant, indicating a discernible temporal pattern in MACEs exemption following surgical procedures.

According to the European Society of Cardiology (ESC) guidelines, the deferral of NCS from drug-eluting stent percutaneous coronary intervention (DES-PCI) for at least six months post-surgery is recommended for ensuring a certain level of cardiovascular event safety[20]. This study corroborates similar outcomes, revealing distinct MACEs occurrence rates of 19% and 2% for two consecutive time intervals centered around the 180-day mark. Not only do these rates exhibit substantial numerical disparities, but they also demonstrate significant statistical differences. This observation lends credence to the notion that the 180-day threshold, equivalent to six months, may represent a meaningful temporal landmark for patients undergoing GCS following coronary stent implantation. Delaying surgery by at least six months appears to confer significant benefits to patients[21–23]. However, previous research has suggested an increased risk of myocardial infarction and cardiovascular death within the first 12 months post-DES-PCI for patients requiring surgery, compared to those without ischemic heart disease[11]. Interestingly, this heightened risk is confined to the initial month post-DES-PCI, suggesting that surgery might be reasonably safe if conducted earlier than the currently recommended timeline[13, 24, 25]. Addressing this complexity, our study meticulously stratifies the post-PCI period within 0-180 days, revealing, as mentioned earlier, notable variations in MACEs occurrence rates not only at the six-month mark but also potentially at the one-month and three-month time points. These findings underscore the importance of considering multiple temporal nodes, beyond the conventional six-month period, when evaluating the occurrence of MACEs post-DES-PCI in patients undergoing GCS.

Roshanov et al. proposed that cancer patients undergoing PCI may necessitate early surgical intervention, potentially increasing the likelihood of emergency surgeries[26]. This implies a challenge in timely cessation of dual antiplatelet therapy or initiation of bridging therapy with intravenous medications before surgical procedures[27–29]. Previous research has predominantly focused on elective surgery patients, and this study addresses this limitation by first confirming, through multifactorial logistic regression analysis, that emergency surgery is indeed an independent risk factor for postoperative MACEs[30]. Subsequently, employing line charts, we compared the daily trends in MACEs occurrence rates between emergency and elective surgeries across the entire time range. The study revealed an overall higher MACEs occurrence rate for emergency surgeries compared to elective surgeries. Moreover, a combined line and density chart illustrated the changing trends in the numbers of emergency and elective surgeries. Notably, in the 0-30 days post-PCI period, the count of emergency surgeries sharply increased, peaking at 28 days, surpassing elective surgeries. Subsequently, elective surgeries gradually increased, eventually surpassing emergency surgeries. This pattern may be attributed to the clinical tendency to postpone surgery within the first month post-PCI for gastrointestinal cancer patients without acute signs of bleeding, obstruction, or tumor rupture, potentially offering greater patient benefits[31]. The heightened propensity for the occurrence of MACEs in the aftermath of GCS underscores the potential elevated risk within a short timeframe after DES-PCI. This observation, to some extent, aligns with the heightened risks associated with surgical interventions conducted within a month, in concordance with the guidelines delineated by the American College of Cardiology[32].

Furthermore, the study delves into the role of the 3-month time point in surgical timing. Park et al. suggested that the surgical-related risk for patients treated with DES-PCI stabilizes after 3-6 months[33]. Barash et al. indicated that early NCS within 3 months of DES implantation may be associated with adverse clinical outcomes[34]. Delaying elective NCS by at least 3 months after DES implantation seems safe and feasible with no significant postoperative adverse cardiac events[35]. By establishing a logistic regression formula linking the variable "Time of surgery" to MACEs, based on the Youden index, the study calculates the time point corresponding to the threshold probability as the 87th day. Additionally, from another statistical perspective, Kaplan-Meier curves were plotted using MACEs exemption rates for each day within the first 30 days postoperatively, and log-rank tests indicated significant statistical differences between the 0-30 days, 30-90 days, and 90-360 days intervals. This further emphasizes that the 3-month mark is a critical time node where significant changes in MACEs occurrence rates occur for patients.

### 4.2 Strength and Limitations

The primary strength of this study lies in its utilization of data from multiple medical centers. By employing a rigorous multi-dimensional statistical approach and controlling for various independent risk factors associated with MACEs, the study stratifies the intervals of surgical timing to substantiate the impact of surgical timing on the outcome of MACEs. However, the study is not without limitations. Firstly, our research adopts a retrospective design, introducing the possibility of selection bias during the case history screening process. Secondly, pertinent data on preoperative antiplatelet therapy regimens and preoperative bridging treatments were not included in our study, potentially exerting an influence on the outcomes of MACEs. Thirdly, the timing of patient surgeries may be influenced by factors within the realm of clinical treatment, such as insufficient discontinuation time of antiplatelet medications and the delay in surgical timing attributable to complications arising from consultations.

### 4.3 Conclusion

The safety for GCS within 6 months after DES-PCI is notably high. In cases where cancer progression is rapid, surgery may be expedited to a window between 3-6 months post-DES implantation. For patients with cancer-related complications necessitating time-sensitive management, surgery can be scheduled earlier, within the period of 1-3 months post-DES implantation. However, the surgical risk is markedly elevated within the first month after DES implantation, and, barring emergencies, surgical procedures should be deferred as much as possible during this early period.

## Data Availability

All data cited in this paper can be provided and backed up to ensure data integrity and traceability. If necessary, the author will provide relevant data and additional information to support the validation and reproduction of the research results. Please contact the author for access permissions and detailed information on the relevant data. We are committed to promoting scientific sharing and transparency to advance research in the field of biotechnology.

https://www.jianguoyun.com/p/DbCNYIcQ1LGsDBj69LIFIAA

## Acknowledgements

We thanked Home for Researcher for language editing service.

## Author contribution statement

Ziyao Xu: Literature review and writing. Xinyu Hao: Data collection and statistical analysis. Jingyang Tian: Data curation. Qiying Song: Supervision. Lei Gao and Tian Li: Resources, Project administration, Writing – review & editing. Xinxin Wang: Resources, Project administration, Writing – review & editing, Funding acquisition.

## Data Availability

https://www.jianguoyun.com/p/DbCNYIcQ1LGsDBj69LIFIAA

## Declarations

### Ethics approval and consent to participate

The authors assert that all procedures contributing to this work comply with the ethical standards of the relevant national and institutional committees on human experimentation and with the Helsinki Declaration of 1975, as revised in 2008. This study received approval from the Research Ethics Committee of the People’s Liberation Army General Hospital, denoted by approval No. S2023-630, and did not require informed patient consent.

### Consent for publication

Patients have provided written inform consent.

## Supplement

**Sfigure1.**
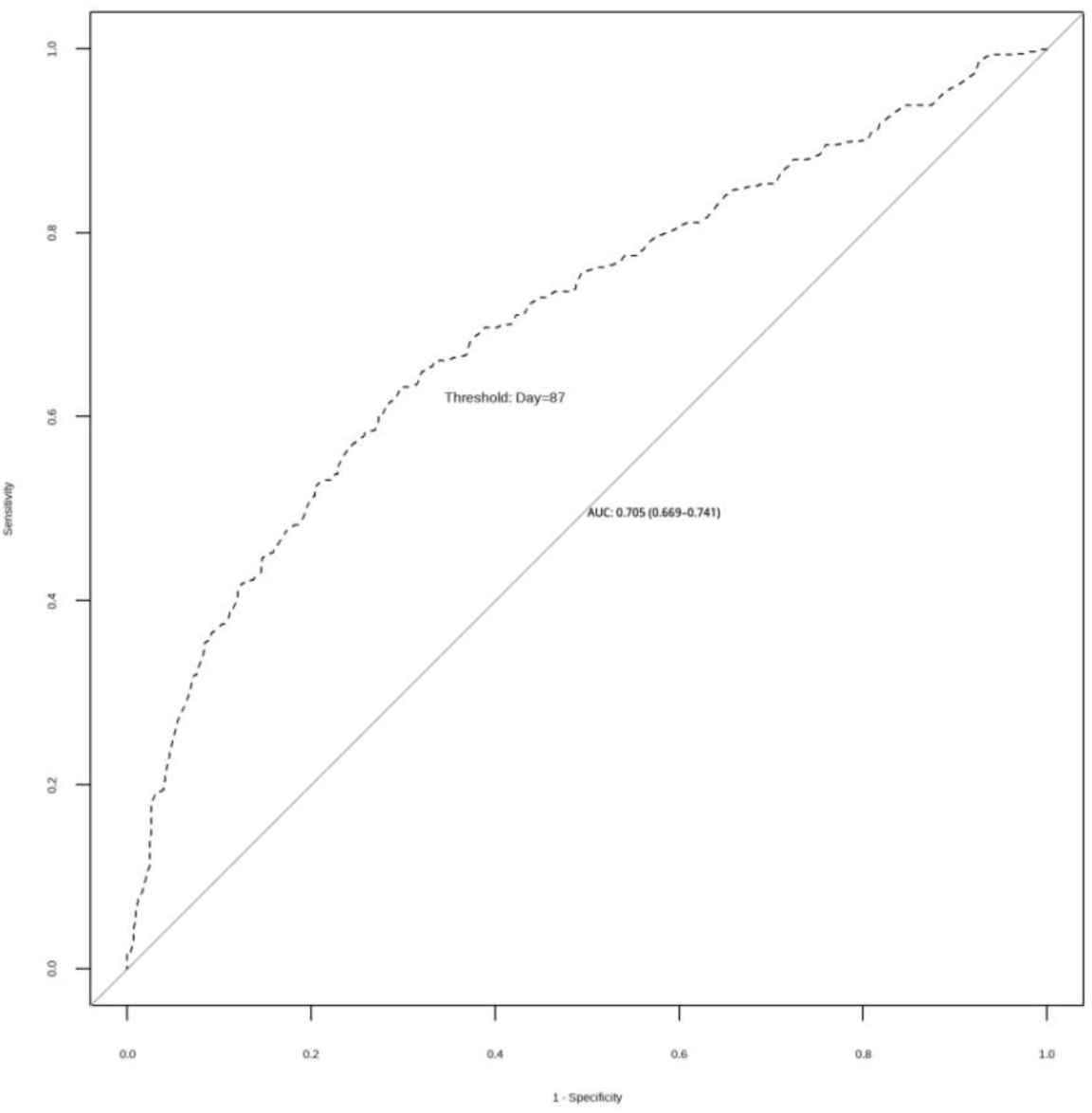
Using ROC curves to determine the probability threshold under the Jordan exponent.

**Sfigure2.**
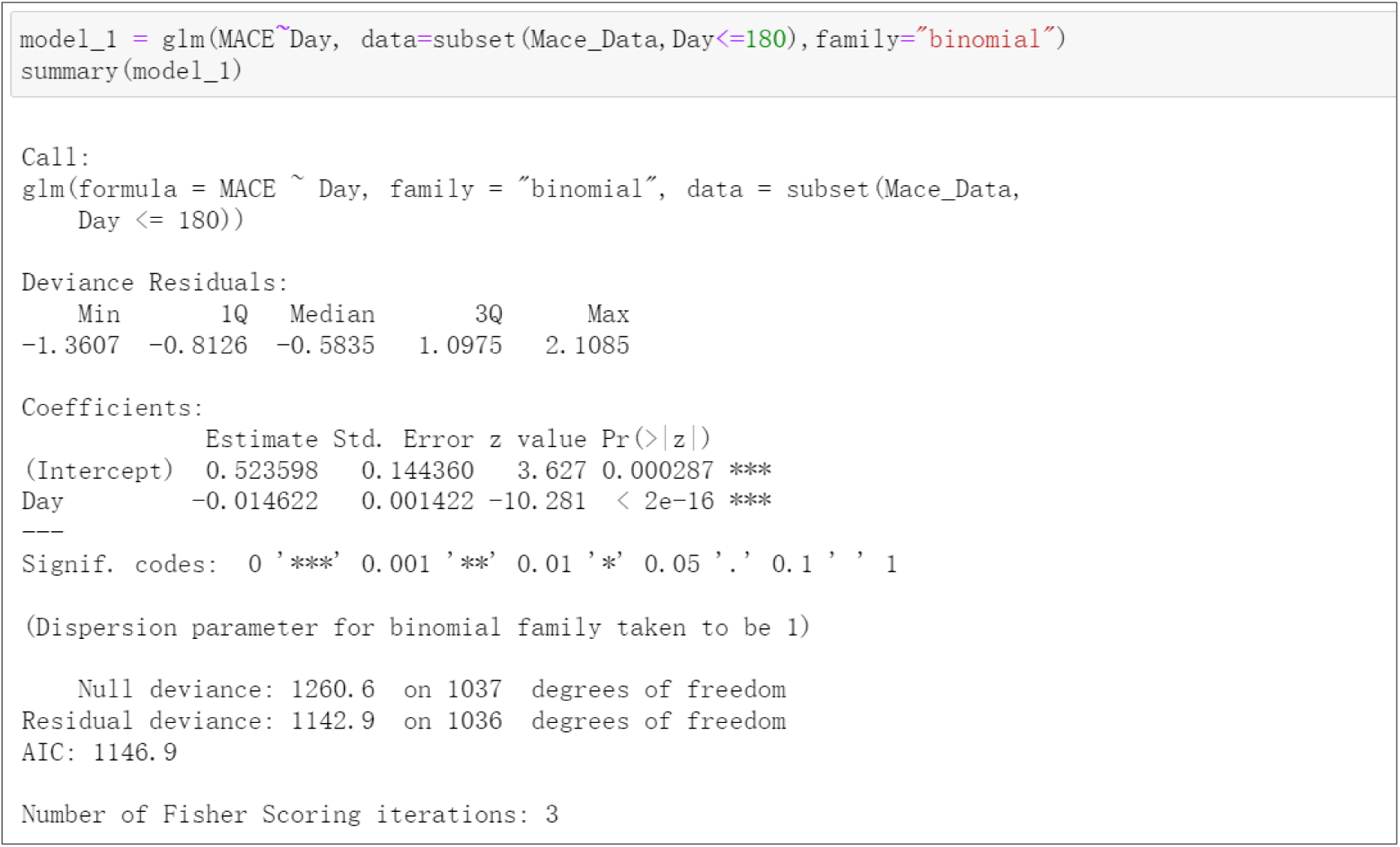
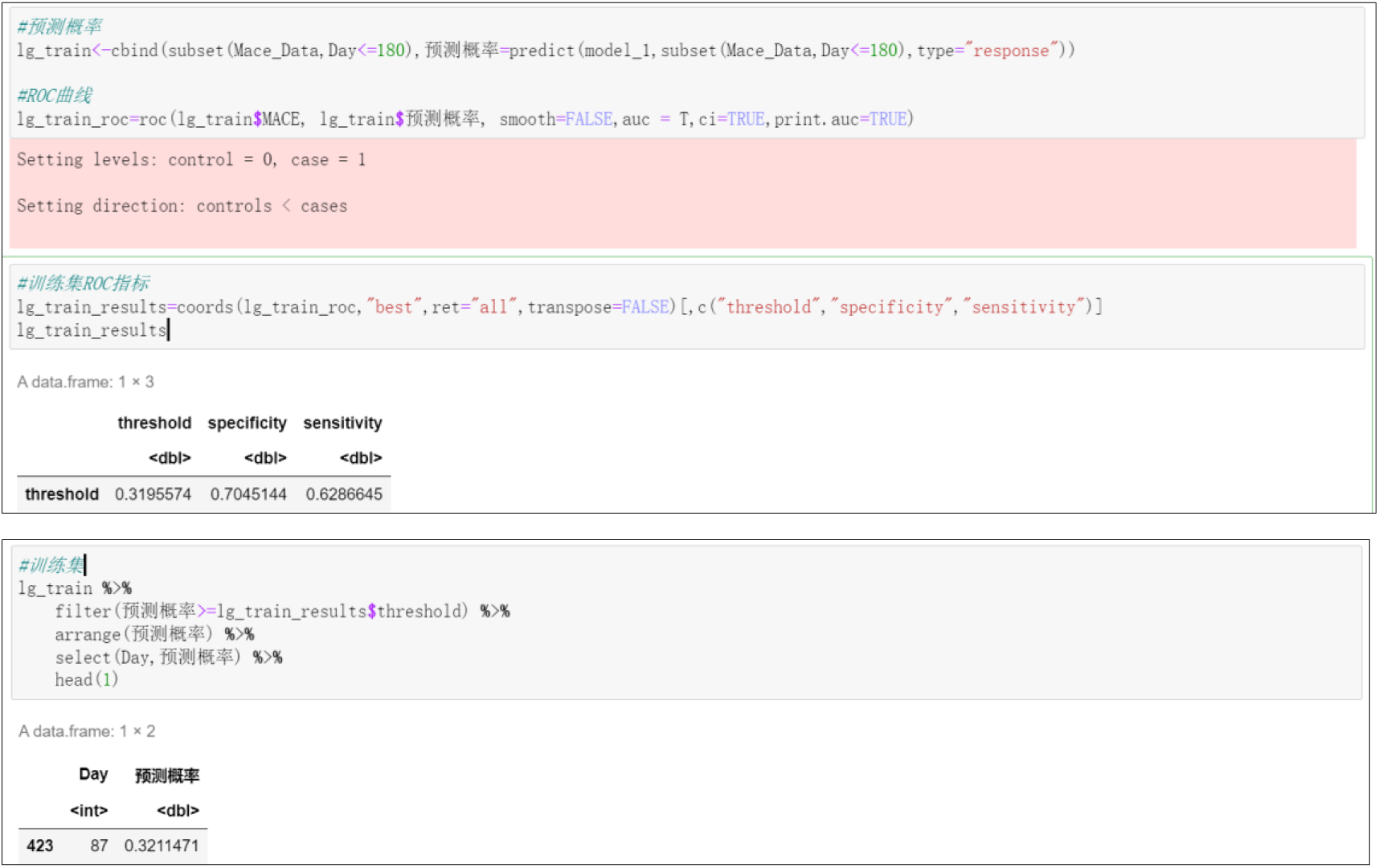
The calculation process of probability threshold under the Jordan index.

